# NLP Framework for Automated Symptom Severity Staging in Heart Failure and COPD Clinical Notes Using Ontology Integration: A Study Protocol

**DOI:** 10.64898/2026.06.27.26356738

**Authors:** Job Inyangala, Fidelis Musena Mukudi, Ronald Ojino, Morris Senghor Shisanya

## Abstract

**Background:** Heart failure (HF) and chronic obstructive pulmonary disease (COPD) are among the leading causes of morbidity and mortality globally, with effective management heavily dependent on accurate severity staging using the New York Heart Association (NYHA) and Global Initiative for Chronic Obstructive Lung Disease (GOLD) classification systems. However, severity information is frequently embedded within unstructured clinical narratives rather than standardized Electronic Health Record (EHR) fields, limiting automated clinical decision support, disease surveillance, and retrospective healthcare analytics. Existing Natural Language Processing (NLP) approaches primarily rely on rule-based keyword extraction or supervised deep learning methods requiring large annotated corpora, which are often unavailable in many healthcare settings. Equally, most current systems inadequately integrate clinical ontologies for semantic reasoning and explainable classification, limiting interoperability and clinical applicability.

**Objective:** This study aims to develop and evaluate an ontology-integrated NLP framework for automated extraction and severity staging of HF and COPD symptoms from de-identified clinical notes using NYHA and GOLD classification systems.

**Methods:** The study will employ a Design Science Research (DSR) methodology to design, implement, and evaluate a hybrid NLP framework integrating rule-based extraction, SNOMED-CT ontology reasoning, and a Bidirectional Long Short-Term Memory with Conditional Random Field (Bi-LSTM-CRF) deep learning architecture for clinical sequence labeling. Approximately 1,000 de-identified clinical notes will be sampled proportionately from publicly available repositories including MIMIC-III/IV, eICU Collaborative Research Database, AmsterdamUMCdb, and MTSamples. Clinical text preprocessing will include tokenization, lemmatization, dependency parsing, abbreviation expansion, and negation detection. Ontology-guided semantic normalization will map extracted symptom entities to standardized SNOMED-CT concepts to support severity staging. Framework performance will be evaluated using precision, recall, F1-score, Cohen’s Kappa, sensitivity, specificity, positive predictive value, negative predictive value, confusion matrices, and correlation analyses against confirmed diagnoses and guideline-based severity classifications.

**Expected Outcomes:** The proposed framework is expected to automate NYHA and GOLD severity staging across heterogeneous clinical note types without reliance on manually annotated severity labels. The ontology-integrated architecture is anticipated to improve semantic consistency, interpretability, and explainability of NLP outputs while enhancing EHR analytics, retrospective clinical audit, and AI-assisted clinical decision support.

**Conclusion:** Findings from this study may provide a scalable and transferable framework for automated severity classification in data-rich but label-poor healthcare environments.

## Introduction

### Background

Heart failure (HF) and chronic obstructive pulmonary disease (COPD) are among the leading causes of morbidity, mortality, and healthcare utilization globally. Cardiovascular diseases account for approximately 17.9 million deaths annually, while chronic respiratory diseases contribute substantially to disability-adjusted life years and reduced quality of life worldwide [1,2]. HF affects more than 64 million people globally and is associated with recurrent hospitalization, reduced functional capacity, and high mortality rates [3]. Similarly, COPD affects over 390 million individuals worldwide and remains a major cause of chronic morbidity and premature mortality, particularly in low- and middle-income countries [4]. The increasing burden of these chronic conditions has intensified the need for efficient systems capable of supporting timely diagnosis, severity staging, risk stratification, and evidence-based clinical decision-making.

Accurate severity staging is central to the management of HF and COPD because treatment decisions, prognostic assessments, and monitoring strategies depend heavily on disease severity classifications. In HF management, the New York Heart Association (NYHA) functional classification categorizes patients into four classes according to symptom burden and physical activity limitation [3]. Similarly, the GOLD framework classifies COPD severity using symptom burden, airflow limitation, and exacerbation risk [4]. These staging systems are clinically important because they guide medication selection, hospitalization decisions, rehabilitation planning, and long-term monitoring. Despite their significance, explicit NYHA and GOLD classifications are inconsistently documented in routine clinical records, with clinicians often describing symptom severity indirectly through narrative clinical notes rather than structured fields [5].

The rapid expansion of Electronic Medical Records (EMRs) has generated massive volumes of unstructured clinical narratives including discharge summaries, physician progress notes, consultation reports, and nursing documentation. These narratives contain rich contextual information regarding patient symptoms, functional limitations, physiological changes, and disease progression [6]. However, much of this information remains inaccessible to automated analytics systems because it is embedded within free-text documentation rather than standardized structured fields. Clinical notes frequently contain abbreviations, synonymous terms, fragmented sentence structures, and institution-specific documentation styles that complicate automated interpretation [7]. Consequently, valuable clinical information related to disease severity is underutilized in clinical audit, disease surveillance, and decision-support systems.

Although structured coding systems such as the International Classification of Diseases (ICD) provide standardized disease categorization, they have important limitations in representing clinical severity and patient functional status. ICD coding systems are primarily designed for billing, epidemiological reporting, and administrative purposes rather than nuanced clinical interpretation [8]. Structured codes rarely capture symptom intensity, temporal progression, or contextual clinical narratives needed for NYHA or GOLD staging. Several studies have demonstrated that relying solely on structured coding significantly underestimates symptom burden compared to Natural Language Processing (NLP)-based extraction from free-text clinical notes [8]. Therefore, there is increasing interest in computational approaches capable of transforming unstructured clinical narratives into structured, clinically meaningful severity information.

### Clinical Severity Frameworks

#### NYHA Classification

The NYHA functional classification is one of the most widely used clinical tools for assessing the severity of HF. The system categorizes patients into four classes based on the degree of limitation imposed by symptoms during physical activity. Class I represents patients with no limitation of physical activity, whereas Class IV includes patients with symptoms even at rest [9]. The NYHA system remains clinically relevant because it correlates strongly with hospitalization risk, mortality, and quality of life outcomes. However, clinicians frequently describe symptoms narratively rather than explicitly assigning NYHA classes, making automated extraction and classification difficult within routine EMRs.

#### GOLD Classification

The GOLD classification system is the standard framework for staging COPD severity. The framework integrates airflow limitation, symptom burden, and exacerbation history to classify patients into severity groups ranging from mild to very severe disease [4]. GOLD staging informs treatment selection, pulmonary rehabilitation planning, and exacerbation prevention strategies. Similar to NYHA staging, clinicians often document COPD severity indirectly through descriptions of dyspnea, oxygen requirements, exercise intolerance, or exacerbation frequency rather than assigning explicit GOLD categories. This creates challenges for automated EHR analytics and clinical decision support systems.

#### Natural Language Processing in Healthcare

Natural Language Processing (NLP) is a branch of artificial intelligence that enables computers to process, interpret, and extract meaningful information from human language (Prasad et al., 2022). In healthcare, NLP has emerged as a critical technology for transforming unstructured clinical text into structured data suitable for research, surveillance, and decision support [10]. NLP applications in healthcare include symptom extraction, medication identification, disease surveillance, adverse event detection, and predictive modeling [6].

Named Entity Recognition (NER) is a foundational NLP task involving the identification and classification of predefined entities such as symptoms, diagnoses, medications, and procedures within clinical narratives [11]. NER systems enable automated extraction of clinically relevant concepts from free-text notes, supporting downstream applications such as disease classification and severity prediction. Recent advances in deep learning architectures, particularly Bidirectional Long Short-Term Memory with Conditional Random Fields (Bi-LSTM-CRF) and transformer-based models, have substantially improved the accuracy of clinical entity recognition [12,13].

Clinical text mining extends beyond entity recognition to include semantic interpretation, relation extraction, and predictive analytics. Deep learning approaches such as recurrent neural networks, convolutional neural networks, and transformer architectures have demonstrated strong performance in extracting clinical information from unstructured EMRs [14]. Nevertheless, many existing NLP systems remain limited to entity detection without translating extracted findings into clinically actionable severity classifications aligned with validated clinical frameworks such as NYHA and GOLD.

#### Ontology-Based Clinical NLP

Clinical ontologies provide structured representations of biomedical knowledge that facilitate semantic interoperability, concept normalization, and explainable clinical reasoning. SNOMED-CT is among the most comprehensive clinical ontology systems and provides standardized terminology for symptoms, diagnoses, procedures, and physiological concepts ([15]. Similarly, the Unified Medical Language System (UMLS) integrates multiple biomedical vocabularies to support concept mapping and semantic consistency across clinical datasets [16].

Ontology integration enhances clinical NLP systems by enabling semantic normalization of synonymous terms, hierarchical reasoning, and contextual interpretation of extracted concepts. Ontology-guided NLP improves semantic interoperability between healthcare systems and facilitates more accurate extraction of clinically meaningful information from heterogeneous clinical narratives [17]. Furthermore, ontology-based approaches improve explainability by linking NLP predictions to standardized clinical concepts and reasoning pathways, thereby reducing the “black-box” limitations associated with many deep learning models [18].

### Existing Gaps

Despite major advances in healthcare NLP, several important gaps remain. Many current systems rely heavily on rule-based methods that depend on manually developed keyword dictionaries and heuristic patterns. Although rule-based systems provide interpretability, they are labor-intensive to maintain and perform poorly when confronted with linguistic variability, abbreviations, or institution-specific documentation practices [19].

Another major limitation is the dependence on large manually annotated datasets required for supervised deep learning models. Annotated severity corpora for HF and COPD are scarce, especially in low-resource healthcare systems, limiting the scalability and transferability of existing models [20]. In addition, many NLP systems demonstrate poor generalizability across institutions due to differences in clinical language, note structures, and documentation practices [21].

Existing studies have also largely focused on single-disease applications or isolated entity recognition tasks rather than integrated severity staging systems capable of simultaneously handling HF and COPD classifications. Few frameworks combine deep learning with ontology-guided semantic reasoning to support explainable and clinically aligned severity staging. Consequently, there remains a need for hybrid NLP architectures that integrate symbolic clinical knowledge with deep sequence learning to achieve scalable, interpretable, and clinically meaningful severity classification.

### Theoretical Framework

#### Frame Semantics Theory

This study is grounded in Frame Semantics Theory proposed by Charles Fillmore, which posits that the meaning of words is understood within structured contextual situations known as semantic frames [22]. Semantic frames represent prototypical situations involving participants, relationships, and contextual roles. In clinical narratives, symptom descriptions activate disease-specific semantic frames associated with severity, progression, physiological limitation, and treatment response.

Predicate-argument structures within Frame Semantics provide a mechanism for representing relationships between symptoms, contextual modifiers, and severity indicators independently of grammatical variation [23]. This theoretical perspective is highly relevant to clinical NLP because clinicians frequently express disease severity indirectly through contextual descriptions rather than explicit staging labels. For example, descriptions such as “shortness of breath at rest” or “unable to climb stairs” activate semantic frames associated with advanced HF severity.

Frame Semantics therefore provides the conceptual foundation for integrating neural pattern recognition with ontology-guided semantic reasoning. Within the proposed framework, semantic frames operationalized through SNOMED-CT mappings and deep learning sequence models will support automated extraction and severity classification of HF and COPD symptoms from unstructured clinical narratives as shown in Figure 1.

**Figure 1.**
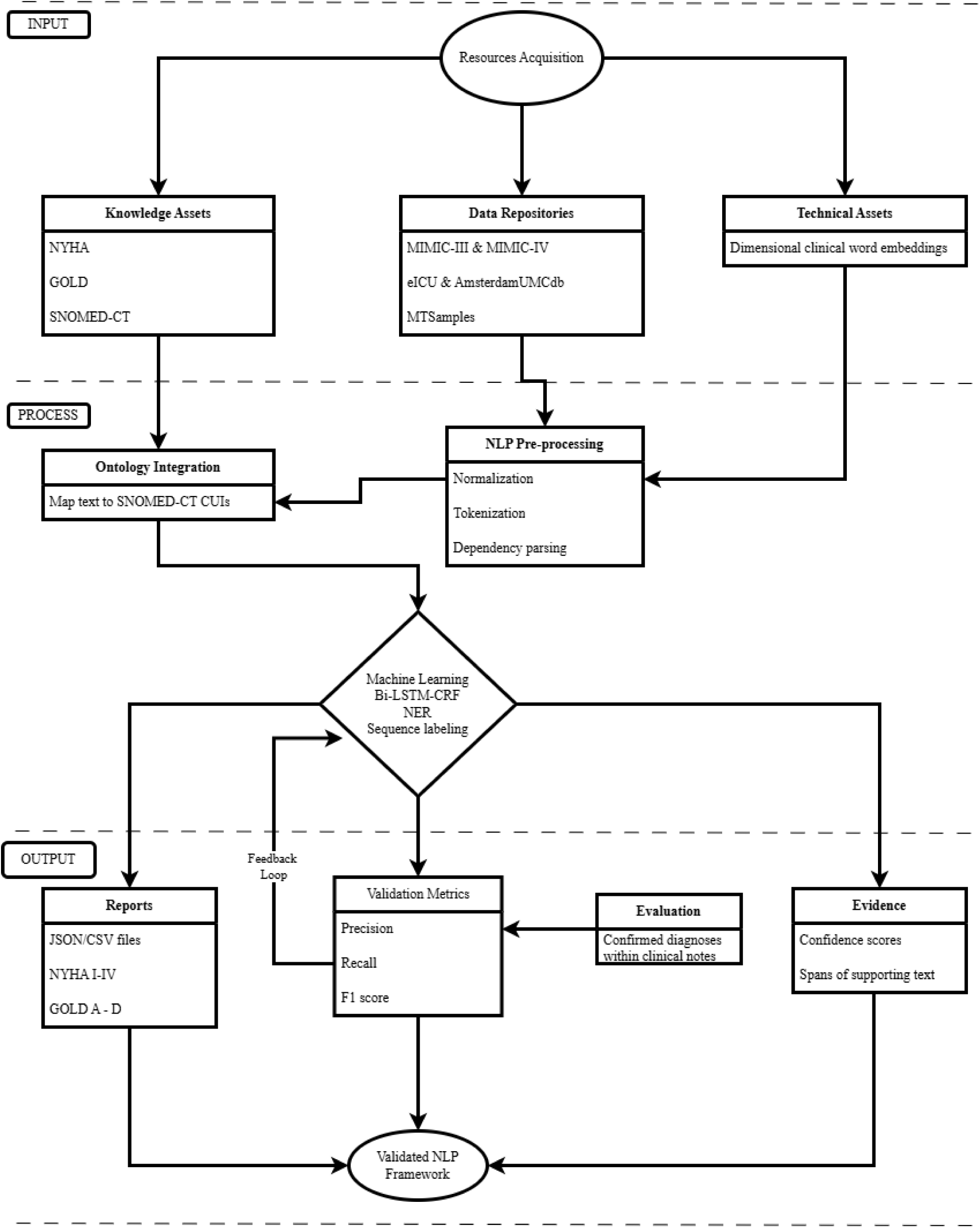
Conceptual Framework.

### Study Aim

The aim of this study is to develop and evaluate an ontology-integrated NLP framework for automated extraction and severity staging of HF and COPD symptoms from de-identified clinical notes using NYHA and GOLD classification systems.

### Study Objectives

#### General Objective

To develop and evaluate an ontology-guided NLP framework for automated severity staging of HF and COPD symptoms from unstructured clinical narratives.

#### Specific Objectives

1. To evaluate the accuracy of entity extraction for HF and COPD symptoms from de-identified clinical notes.
2. To assess ontology-guided severity classification of HF and COPD symptoms using NYHA and GOLD staging frameworks.
3. To evaluate the performance of the NLP framework against confirmed diagnoses within clinical records.

## Materials And Methods

### Study Design

This study will adopt a Design Science Research (DSR) methodology to develop and evaluate an ontology-integrated Natural Language Processing (NLP) framework for automated severity staging of heart failure (HF) and chronic obstructive pulmonary disease (COPD) from unstructured clinical narratives. Design Science Research is an established methodological approach that focuses on the systematic development, implementation, and evaluation of innovative technological artefacts designed to address complex real-world problems [24]. In healthcare informatics, DSR is particularly appropriate for studies involving development of artificial intelligence systems, clinical decision-support frameworks, and computational models intended for practical implementation and evaluation.

The proposed framework will integrate rule-based NLP, ontology-guided semantic reasoning, and deep learning architectures to automate extraction and staging of HF and COPD symptoms using validated clinical severity frameworks, specifically the New York Heart Association (NYHA) and Global Initiative for Chronic Obstructive Lung Disease (GOLD) classification systems. The study will therefore focus not only on technical model development but also on demonstrating clinical interpretability, semantic interoperability, and applicability within Electronic Health Record (EHR) analytics.

The study will be implemented in three interrelated phases. Phase I will involve conceptual framework design and architecture specification. During this phase, the overall NLP pipeline structure, ontology integration strategy, and deep learning workflow will be defined based on identified gaps in clinical NLP literature. The architecture will specify interactions between preprocessing modules, ontology reasoning layers, and severity classification mechanisms.

Phase II will involve framework development and implementation. This phase will include data acquisition, preprocessing of clinical notes, ontology mapping using SNOMED-CT terminology resources, and training of the Bidirectional Long Short-Term Memory with Conditional Random Field (Bi-LSTM-CRF) model for clinical entity recognition. The framework will incorporate semantic normalization, negation detection, and severity inference logic for NYHA and GOLD staging.

Phase III will involve framework validation and evaluation. Framework performance will be assessed using standard NLP evaluation metrics including precision, recall, F1-score, sensitivity, specificity, positive predictive value (PPV), negative predictive value (NPV), confusion matrices, and Cohen’s Kappa agreement measures. The framework outputs will further be evaluated against confirmed diagnoses and clinically expected severity distributions within the selected datasets.

The DSR methodology is appropriate for this study because it supports iterative refinement, integration of theoretical and technical components, and rigorous evaluation of system performance within realistic clinical contexts.

### Study Setting and Data Sources

The study will utilize publicly available de-identified clinical datasets containing unstructured Electronic Medical Record (EMR) narratives relevant to HF and COPD. These repositories contain discharge summaries, physician notes, progress reports, nursing documentation, and intensive care records suitable for NLP-based severity staging research. The datasets were selected because of their accessibility, clinical diversity, structured metadata, and widespread use in healthcare NLP research.

### MIMIC-III

The Medical Information Mart for Intensive Care III (MIMIC-III) is a large publicly accessible critical care database developed by the Massachusetts Institute of Technology (MIT) in collaboration with Beth Israel Deaconess Medical Center, USA. The database contains de-identified clinical data from over 40,000 intensive care admissions collected between 2001 and 2012 [25]. MIMIC-III includes discharge summaries, nursing notes, physician documentation, laboratory records, medication information, and ICD-9 diagnostic codes. For this study, HF- and COPD-related clinical notes will be retrieved using diagnosis-linked SQL queries through authenticated Google BigQuery access.

### MIMIC-IV

MIMIC-IV is the updated version of the MIMIC database and contains de-identified healthcare data for over 70,000 ICU admissions collected between 2008 and 2019 [26]. The database incorporates improved data organization, ICD-10 coding structures, and expanded clinical note repositories. MIMIC-IV will provide contemporary clinical narratives for evaluating framework performance across modern EHR documentation systems.

### eICU Collaborative Research Database

The eICU Collaborative Research Database is a multicenter intensive care database containing de-identified health data associated with approximately 200,000 ICU admissions across multiple hospitals in the United States [27]. The database includes physician notes, nursing notes, treatment plans, medication records, and diagnosis-linked clinical documentation. The multicenter structure of eICU provides linguistic and institutional variability that is valuable for assessing framework generalizability across healthcare settings.

### AmsterdamUMCdb

AmsterdamUMCdb is a European intensive care database developed by Amsterdam University Medical Centers containing de-identified clinical data from approximately 23,000 ICU admissions [28]. The database includes clinical narratives, physiological observations, treatment documentation, and admission summaries. Inclusion of AmsterdamUMCdb will improve external dataset diversity and support cross-regional validation of ontology-guided severity staging.

### MTSamples

MTSamples is a publicly available corpus of approximately 5,000 medical transcription reports across multiple clinical specialties. The repository contains outpatient consultations, operative reports, discharge summaries, and specialty-specific medical narratives. MTSamples is widely used in healthcare NLP research because it contains heterogeneous clinical language and varied documentation styles. The dataset will be extracted through automated text retrieval in accordance with repository usage terms [29].

### Eligibility Criteria

#### Inclusion Criteria

Clinical notes will be eligible for inclusion if they meet the following criteria:

1. Unstructured English-language clinical narratives including discharge summaries, physician progress notes, consultation reports, and nursing documentation.
2. Clinical notes containing symptomatic descriptions relevant to HF or COPD severity staging, including dyspnea, fatigue, exercise intolerance, edema, cough, wheezing, oxygen dependence, or respiratory distress.
3. Clinical records associated with adult patients aged 18 years and above.
4. Notes linked to confirmed HF or COPD diagnoses using ICD-9, ICD-10, or documented clinical diagnosis.
5. Notes originating from the selected publicly accessible repositories: MIMIC-III, MIMIC-IV, eICU, AmsterdamUMCdb, and MTSamples.

#### Exclusion Criteria

Clinical notes will be excluded if they meet any of the following criteria:

1. Non-English clinical narratives.
2. Notes lacking sufficient descriptive symptom information relevant to NYHA or GOLD staging.
3. Duplicate clinical records.
4. Highly redacted or incomplete notes lacking contextual clinical meaning.
5. Structured records containing only ICD codes without accompanying free-text narratives.
6. Pediatric patient records involving individuals younger than 18 years.
7. Notes unrelated to HF or COPD conditions.

### Sample Size and Sampling Strategy

#### Population Estimates

The study will utilize an estimated eligible population of approximately 54,000 clinical notes obtained from the five selected repositories. The estimated population distribution is as follows:

- MTSamples: approximately 5,000 notes
- MIMIC-III/IV: approximately 27,000 notes
- eICU Collaborative Research Database: approximately 18,000 notes
- AmsterdamUMCdb: approximately 4,000 notes

These repositories collectively provide sufficient clinical diversity, note heterogeneity, and linguistic variation necessary for robust NLP framework development and evaluation.

#### Sampling Strategy

A proportionate stratified random sampling strategy will be used to ensure representative inclusion of clinical notes from each data source. Stratification will be based on dataset origin, with each repository considered a separate sampling stratum. This approach will preserve dataset diversity while minimizing overrepresentation from larger repositories.

Within each stratum, eligible notes meeting inclusion criteria will be assigned unique identifiers and selected using computer-generated random sampling procedures. Stratified sampling is appropriate because the selected repositories differ substantially in note structure, clinical documentation style, and institutional characteristics. The approach therefore enhances generalizability of the framework across heterogeneous clinical environments.

#### Sample Size Determination

Sample size estimation will be based on Cochran’s formula for large populations:

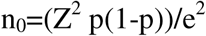

Where:

- n_0_= required sample size
- Z= standard normal deviate corresponding to 95% confidence level (1.96)
- p= estimated proportion (0.5 used to maximize variability)
- e= margin of error

Using a 95% confidence level and a margin of error of approximately 3%, the estimated samplesize is approximately 1,000 clinical notes. This sample size is considered adequate for NLP framework training, validation, and evaluation while maintaining computational feasibility and will be proportionately allocated across four databases as shown in Table 1.

**Table 1:** Sample Allocation Table.

| Database | Estimated Population | Population Proportion | Allocated Sample |
| --- | --- | --- | --- |
| MTSamples | 5,000 | 9.26% | 93 |
| MIMIC-III/IV | 27,000 | 50.00% | 500 |
| eICU Collaborative Research Database | 18,000 | 33.33% | 333 |
| AmsterdamUMCdb | 4,000 | 7.41% | 74 |
| **Total** | **54,000** | **100%** | **1,000** |

The final sample will therefore consist of approximately 1,000 de-identified clinical notes proportionately distributed across the selected repositories to support balanced framework development and validation.

### NLP Framework Development

#### Overall Framework Architecture

The proposed Natural Language Processing (NLP) framework will consist of a multi-layer hybrid architecture integrating clinical text preprocessing, ontology-guided semantic reasoning, rule-based severity inference, and deep learning sequence labeling. The framework is designed to automate extraction and severity staging of heart failure (HF) and chronic obstructive pulmonary disease (COPD) symptoms from unstructured clinical narratives using validated New York Heart Association (NYHA) and Global Initiative for Chronic Obstructive Lung Disease (GOLD) classification systems.

The architecture combines symbolic reasoning and statistical learning approaches to address limitations associated with standalone rule-based or purely deep learning NLP systems. The framework will integrate domain-specific ontology resources such as Systematized Nomenclature of Medicine–Clinical Terms (SNOMED-CT) with Bidirectional Long Short-Term Memory with Conditional Random Field (Bi-LSTM-CRF) sequence models to improve semantic consistency, interpretability, and generalizability across heterogeneous clinical datasets [17,30]. The framework will operate through four sequential processing stages: (1) data acquisition and preprocessing, (2) ontology integration and semantic normalization, (3) rule-based severity classification, and (4) deep learning-based sequence labeling and entity recognition.

#### Stage 1: Data Acquisition and Preprocessing

The first stage of the framework will involve acquisition, cleaning, and preprocessing of de-identified clinical narratives obtained from MIMIC-III, MIMIC-IV, eICU Collaborative Research Database, AmsterdamUMCdb, and MTSamples repositories. The preprocessing pipeline is intended to standardize heterogeneous clinical text formats and improve downstream NLP performance.

#### Cleaning

Initial text cleaning procedures will involve removal of duplicate entries, irrelevant metadata, malformed characters, excessive whitespace, HTML tags, and formatting inconsistencies. Placeholder identifiers generated during de-identification processes will be normalized to preserve syntactic consistency without introducing patient-identifiable information. Clinical abbreviations commonly encountered in HF and COPD documentation, such as “SOB” for shortness of breath and “CHF” for congestive heart failure, will be expanded using domain-specific abbreviation dictionaries.

#### Tokenization

Tokenization will involve segmentation of clinical narratives into individual lexical units or tokens. Sentence boundary detection and token segmentation will be performed using spaCy and MedSpaCy libraries optimized for clinical text processing [31]. Tokenization is essential for downstream sequence labeling because it enables contextual representation of clinical entities such as symptoms, physiological findings, temporal expressions, and severity descriptors.

#### Lemmatization

Lemmatization will be used to reduce words to their canonical root forms while preserving semantic meaning. This process minimizes lexical variability by standardizing inflected terms such as “breathing,” “breathed,” and “breathes” into a common base form. Lemmatization improves semantic consistency and facilitates ontology matching during concept normalization [32].

#### Part-of-Speech (POS) Tagging

Part-of-speech tagging will assign grammatical labels such as nouns, verbs, adjectives, and adverbs to individual tokens within clinical narratives. POS tagging assists in identifying symptom descriptors, severity modifiers, and contextual qualifiers. Clinical NLP studies have demonstrated that POS tagging improves entity extraction performance by enabling syntactic differentiation between disease mentions and contextual descriptions [13].

#### Dependency Parsing

Dependency parsing will be applied to identify syntactic relationships between words within clinical sentences. The parser will determine grammatical dependencies between symptom mentions, severity modifiers, negation terms, and temporal qualifiers. Dependency parsing is particularly important in clinical narratives where disease severity may be expressed through complex sentence structures. For example, phrases such as “progressive dyspnea on minimal exertion” require contextual linkage between symptoms and activity limitations for accurate severity interpretation.

#### Stage 2: Ontology Integration

The second stage of the framework will focus on ontology-guided semantic normalization and concept mapping using SNOMED-CT terminology resources. Ontology integration is essential for improving semantic interoperability, reducing lexical ambiguity, and enabling explainable clinical reasoning [15].

#### SNOMED-CT Mapping

Extracted clinical entities will be mapped to standardized SNOMED-CT concepts using ontology lookup mechanisms. SNOMED-CT provides hierarchical biomedical terminology covering symptoms, diagnoses, procedures, physiological findings, and clinical states. Concept mapping will allow synonymous expressions such as “shortness of breath,” “dyspnea,” and “breathlessness” to be normalized into semantically equivalent standardized concepts.

#### Semantic Normalization

Semantic normalization will involve transforming heterogeneous clinical expressions into consistent ontology-linked representations. The process will improve interoperability across datasets and reduce variability caused by institution-specific documentation styles. Semantic normalization is critical because clinicians frequently describe similar clinical phenomena using different lexical expressions [16].

#### Fuzzy Matching

Fuzzy matching algorithms based on Levenshtein distance and cosine similarity measures will be implemented to identify semantically similar concepts despite spelling variations, abbreviations, or typographical inconsistencies. Fuzzy matching is necessary because clinical narratives commonly contain misspellings, shorthand notations, and informal documentation patterns that may not exactly correspond to ontology dictionary entries.

#### Negation Detection

Negation detection mechanisms will be incorporated to distinguish affirmed symptoms from negated clinical findings. Rule-based contextual algorithms similar to NegEx and ConText approaches will identify phrases such as “denies dyspnea,” “no wheezing,” or “without edema” and prevent false-positive symptom extraction. Negation detection is essential for ensuring clinical validity of extracted symptom entities and downstream severity classification [33,34].

#### Stage 3: Rule-Based Severity Classification

The third stage of the framework will implement rule-based severity classification aligned with validated NYHA and GOLD staging criteria. This component will combine explicit keyword detection with ontology-guided reasoning to infer disease severity from symptom narratives.

#### NYHA Rule Engine

The NYHA rule engine will classify HF severity into Classes I–IV based on symptom burden and functional limitation described in clinical notes. The engine will use weighted keyword dictionaries containing symptom descriptors such as “dyspnea on exertion,” “fatigue,” “orthopnea,” “exercise intolerance,” and “symptoms at rest.” Contextual severity indicators will be mapped to NYHA stages according to established clinical criteria.

#### GOLD Rule Engine

The GOLD rule engine will classify COPD severity using symptom burden, airflow limitation indicators, oxygen dependence, and exacerbation-related descriptions. Clinical expressions such as “frequent exacerbations,” “oxygen therapy,” “chronic cough,” and “severe wheezing” will be weighted according to GOLD severity thresholds. Where available, spirometry-related expressions such as FEV1 percentages will contribute additional severity evidence.

#### Severity Weighting

The framework will implement severity weighting mechanisms to quantify relative contributions of extracted clinical entities toward final severity classification. High-severity indicators such as “respiratory distress,” “symptoms at rest,” and “end-stage disease” will receive stronger weights than moderate or mild symptom descriptors. Weighted scoring will allow aggregation of symptom evidence across heterogeneous clinical narratives.

#### Ontology Fallback Logic

Ontology fallback logic will be implemented for clinical notes lacking explicit NYHA or GOLD terminology. In such cases, ontology-linked symptom entities and severity modifiers will be used to infer probable severity categories through semantic reasoning. For example, ontology-linked concepts related to functional limitation and oxygen dependence may infer advanced HF or COPD severity even in the absence of explicit staging labels. This hybrid approach improves framework coverage and reduces dependence on explicitly documented severity classifications.

#### Stage 4: Deep Learning Component

The final stage of the framework will involve implementation of a Bidirectional Long Short-Term Memory with Conditional Random Field (Bi-LSTM-CRF) architecture for sequence labeling and clinical entity recognition.

#### Bi-LSTM-CRF Architecture

The Bi-LSTM-CRF model combines bidirectional recurrent neural networks with structured probabilistic decoding for clinical sequence labeling tasks. Bidirectional Long Short-Term Memory layers capture contextual dependencies from both preceding and succeeding words within clinical narratives, while the Conditional Random Field layer models valid label transitions across token sequences [35]. This architecture has demonstrated strong performance in healthcare NLP tasks involving Named Entity Recognition (NER).

The model architecture will include:

- token embedding layers,
- bidirectional LSTM layers,
- linear projection layers,
- and CRF decoding layers.

Clinical word embeddings pretrained on biomedical corpora will be used to improve contextual representation of domain-specific terminology.

#### Sequence Labeling

Sequence labeling will be performed using BIO (Beginning–Inside–Outside) annotation schemes to identify and classify clinical entities including:

- symptoms,
- severity indicators,
- physiological measures,
- and temporal expressions.

The sequence model will learn contextual relationships between symptom descriptors and severity modifiers within clinical narratives.

#### Auto-Annotation Strategy

Because manually annotated severity corpora are limited, the study will employ an ontology-guided auto-annotation strategy. SNOMED-CT mappings and rule-based severity inference outputs generated during earlier stages will provide weak supervision labels for training the Bi-LSTM-CRF model. This approach reduces dependence on labor-intensive manual annotation while enabling scalable model development across large clinical datasets.

#### Training-Validation-Test Split

The dataset will be divided into training, validation, and testing subsets using a 70:15:15 split ratio. The training set will be used for model learning, the validation set for hyperparameter tuning and early stopping, and the testing set for final performance evaluation. Stratified splitting procedures will ensure balanced representation of HF and COPD notes across all datasets.

Model optimization will include:

- early stopping,
- dropout regularization,
- OneCycle learning rate scheduling,
- and weighted loss functions to address class imbalance.

Framework performance will ultimately be evaluated using precision, recall, F1-score, sensitivity, specificity, confusion matrices, and agreement analyses against confirmed diagnoses and ontology-guided severity classifications.

### Model Training and Optimization

#### Training Pipeline

The deep learning component of the proposed Natural Language Processing (NLP) framework will be trained using a supervised learning approach to enable accurate extraction and classification of clinical entities associated with heart failure (HF) and chronic obstructive pulmonary disease (COPD). The training pipeline is designed to optimize model performance while minimizing overfitting, class imbalance, and noise commonly associated with clinical text datasets.

#### Supervised Learning

The Bidirectional Long Short-Term Memory with Conditional Random Field (Bi-LSTM-CRF) architecture will be trained using supervised sequence labeling techniques. In supervised learning, the model learns associations between input clinical text sequences and corresponding target labels derived from ontology-guided auto-annotation processes. Clinical entities will be represented using BIO (Beginning–Inside–Outside) tagging schemes to identify symptom entities, severity descriptors, physiological measures, and temporal expressions within unstructured clinical narratives.

The auto-annotation strategy developed during ontology integration will generate weak supervision labels using SNOMED-CT concept mappings and rule-based severity inference outputs. These labels will provide structured training targets for the sequence labeling model while reducing reliance on labor-intensive manual annotation. The training process will involve iterative optimization of model parameters to minimize prediction error between generated labels and predicted outputs.

Clinical text sequences will first be transformed into vector representations using pretrained biomedical word embeddings to preserve semantic relationships among medical concepts. The Bi-LSTM layers will then process token sequences bidirectionally, enabling the model to capture contextual dependencies from both preceding and succeeding words within clinical narratives. Finally, the Conditional Random Field (CRF) layer will optimize valid label transitions across sequences, improving consistency of predicted entity labels [36].

#### Hyperparameter Tuning

Hyperparameter tuning will be conducted to optimize model performance and improve generalizability across heterogeneous clinical datasets. Important hyperparameters to be optimized will include:

- learning rate,
- batch size,
- embedding dimensions,
- hidden layer size,
- dropout probability,
- optimizer selection,
- and number of training epochs.

A systematic search strategy combining grid search and validation-based optimization will be applied using the validation dataset. Hyperparameter combinations yielding the highest weighted F1-score and lowest validation loss will be selected for final model implementation. Model tuning is important because inappropriate hyperparameter configurations can lead to unstable convergence, poor generalization, or excessive computational complexity [14].

#### Early Stopping

Early stopping mechanisms will be implemented to prevent overfitting during model training. During iterative training, model performance on the validation dataset will be monitored continuously after each training epoch. Training will terminate automatically when validation loss fails to improve over a predefined number of consecutive epochs.

Early stopping is particularly important in healthcare NLP because clinical datasets often contain imbalanced entity distributions and noisy annotations that increase the risk of memorization rather than true generalization [13]. By halting training once performance stabilizes, the framework will improve robustness and reduce unnecessary computational overhead.

#### Regularization

Regularization techniques will be incorporated to improve model generalizability and reduce overfitting. The model will utilize L2 weight regularization to penalize excessively large parameter values during optimization. Regularization constrains model complexity and encourages the learning of generalized linguistic patterns rather than dataset-specific noise. In addition, class-weighted loss functions will be implemented to address imbalance among entity categories. Clinical narratives often contain disproportionate representation of common symptom entities relative to rare severity indicators. Weighted loss functions will therefore improve sensitivity toward minority entity classes critical for accurate severity staging.

### Optimization Techniques

#### OneCycleLR Scheduling

The training process will implement the OneCycle Learning Rate (OneCycleLR) scheduling strategy to improve optimization efficiency and convergence stability. OneCycleLR dynamically adjusts the learning rate during training by initially increasing it to accelerate exploration of parameter space before gradually reducing it to facilitate stable convergence [37].

This strategy has demonstrated strong performance in deep learning optimization because it allows faster convergence while reducing the risk of local minima entrapment. In clinical NLP applications, adaptive learning rate scheduling improves model stability when processing heterogeneous and noisy textual datasets. The OneCycleLR approach is therefore expected to improve training efficiency and final predictive performance of the Bi-LSTM-CRF architecture.

#### Dropout

Dropout regularization will be applied within embedding and hidden layers of the Bi-LSTM architecture to reduce overfitting and improve model robustness. During training, dropout randomly deactivates a proportion of neurons within the network, preventing excessive co-adaptation among features [38].

The dropout mechanism encourages the model to learn distributed and generalized representations of clinical language patterns rather than memorizing specific textual sequences. A dropout probability ranging between 0.2 and 0.5 will be evaluated during hyperparameter tuning to determine the optimal balance between regularization strength and predictive performance.

#### Label-Noise Regularization

Because the framework relies partly on ontology-guided auto-annotation rather than fully manually curated labels, the training data may contain weak supervision errors and noisy entity annotations. Label-noise regularization techniques will therefore be implemented to reduce sensitivity to mislabeled or uncertain training examples.

Noise-robust training approaches including confidence-based weighting and smoothed target distributions will be used to minimize propagation of annotation errors during optimization. These methods improve resilience of the deep learning model when learning from automatically generated labels and have been shown to enhance generalizability in weakly supervised NLP systems [39].

Collectively, these optimization strategies are intended to improve training stability, reduce overfitting, enhance robustness to noisy annotations, and maximize the accuracy and generalizability of the ontology-integrated NLP framework across diverse clinical text repositories.

## Outcome Measures

### Primary Outcomes

#### Entity Extraction Performance

The primary outcome of this study will be the performance of the Natural Language Processing (NLP) framework in accurately extracting clinically relevant entities associated with heart failure (HF) and chronic obstructive pulmonary disease (COPD) from unstructured clinical narratives. Extracted entities will include:

- symptoms,
- severity descriptors,
- physiological measurements,
- temporal expressions,
- and functional limitation indicators.

Examples of target entities include dyspnea, fatigue, orthopnea, wheezing, exercise intolerance, oxygen dependence, edema, exacerbation history, and airflow limitation descriptions. Entity extraction performance is important because accurate recognition of symptom-related concepts forms the foundation for downstream ontology-guided severity classification and diagnostic inference.

The performance of the entity extraction component will be evaluated using standard Named Entity Recognition (NER) metrics including precision, recall, and F1-score across all entity categories. High extraction performance will indicate the framework’s ability to reliably identify clinically meaningful concepts from heterogeneous Electronic Health Record (EHR) narratives.

#### Severity Classification Accuracy

The second primary outcome will be the accuracy of automated severity classification for HF and COPD using New York Heart Association (NYHA) and Global Initiative for Chronic Obstructive Lung Disease (GOLD) staging systems, respectively. Severity classification performance will measure the framework’s ability to correctly assign:

- NYHA Classes I–IV for HF,
- and GOLD severity categories for COPD,

based on symptom narratives and ontology-guided semantic reasoning.

Classification accuracy will be assessed by comparing predicted severity stages against guideline-consistent staging logic and diagnosis-linked clinical documentation. Accurate severity classification will demonstrate the framework’s potential utility in automated clinical decision support, retrospective audit, and disease surveillance applications.

### Secondary Outcomes

#### Ontology Agreement

Ontology agreement will evaluate consistency between rule-based severity inference and ontology-guided semantic classification pathways within the framework. Agreement analysis will determine the extent to which SNOMED-CT semantic reasoning produces severity classifications aligned with explicit rule-engine outputs.

High agreement between ontology reasoning and rule-based inference will indicate semantic consistency and reliability of the hybrid framework architecture. This outcome is particularly important because ontology integration is intended to improve interpretability and reduce variability associated with heterogeneous clinical language.

#### Diagnostic Correlation

Diagnostic correlation will assess the relationship between predicted severity classifications and confirmed HF or COPD diagnoses within the selected datasets. The framework will be evaluated for its ability to appropriately route HF narratives toward NYHA staging and COPD narratives toward GOLD classification systems.

Strong diagnostic correlation will indicate clinical validity of the framework and demonstrate alignment between NLP-derived severity outputs and underlying disease diagnoses.

#### Framework Interpretability

Framework interpretability will assess the extent to which severity predictions can be explained through identifiable clinical entities, ontology mappings, and semantic reasoning pathways. Unlike black-box deep learning systems, the proposed hybrid architecture integrates symbolic ontology reasoning to improve transparency and explainability of predictions.

Interpretability assessment will focus on:

- traceability of extracted entities,
- ontology-linked severity mappings,
- and explainable severity inference pathways.

This outcome is important for promoting clinician trust, regulatory acceptance, and future integration into clinical decision-support environments.

### Data Analysis

Quantitative and qualitative analytical approaches will be used to evaluate framework performance across the three study objectives. Statistical analyses will be conducted using Python-based libraries including scikit-learn, PyTorch, spaCy, pandas, NumPy, and SciPy. Evaluation procedures will focus on entity extraction performance, ontology-guided severity classification, and diagnostic validity.

### Objective 1: Entity Extraction Accuracy

The first objective will assess the accuracy of the NLP framework in extracting clinically relevant HF and COPD entities from unstructured clinical narratives.

#### Metrics

##### Precision

Precision will measure the proportion of correctly identified entities among all entities predicted by the framework. High precision indicates a low false-positive rate and reflects the framework’s ability to avoid incorrect entity predictions.

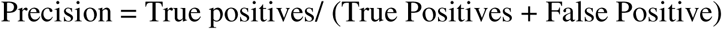

##### Recall

Recall will measure the proportion of true clinical entities successfully identified by the framework among all actual entities present within the dataset. High recall reflects the model’s sensitivity in detecting clinically relevant information.

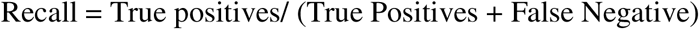

##### F1-score

The F1-score will provide a harmonic mean of precision and recall, offering a balanced measure of extraction performance across entity categories.

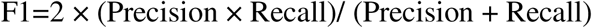

Weighted and macro-averaged F1-scores will be calculated to account for class imbalance across symptom and severity entity categories.

##### Error Analysis

Qualitative error analysis will be conducted to identify major sources of extraction failure and improve interpretability of model performance.

##### False Positives

False-positive analysis will examine instances where the framework incorrectly identifies non-clinical text or irrelevant concepts as symptom entities. These errors may arise from ambiguous terminology, contextual misunderstanding, or abbreviation confusion.

##### False Negatives

False-negative analysis will assess clinically relevant entities missed by the framework. Particular attention will be given to complex symptom descriptions, implicit severity narratives, and institution-specific terminology.

##### Abbreviation Handling

Because clinical narratives frequently contain abbreviations and shorthand notations, targeted analysis will assess the effectiveness of abbreviation expansion and normalization strategies. Performance differences before and after abbreviation normalization will be examined to determine their impact on extraction accuracy.

### Objective 2: Ontology-Guided Severity Classification

The second objective will evaluate ontology-guided severity classification using NYHA and GOLD staging systems.

#### Agreement Analysis

##### Cohen’s Kappa

Cohen’s Kappa statistic will be used to measure agreement between:

- rule-based severity classification,
- and ontology-guided semantic classification outputs.

Cohen’s Kappa adjusts for agreement occurring by chance and is appropriate for evaluating consistency between independent classification methods.

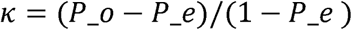

Where:

- P_orepresents observed agreement,
- and P_erepresents expected agreement by chance.

Interpretation of Kappa values will follow established guidelines:

- <0.20: slight agreement,
- 0.21–0.40: fair agreement,
- 0.41–0.60: moderate agreement,
- 0.61–0.80: substantial agreement,
- 0.80: near-perfect agreement.

##### Macro-F1

Macro-averaged F1-scores will be used to evaluate balanced classification performance across all NYHA and GOLD severity categories. Macro-F1 is particularly important because severity stages may not be equally represented within the datasets.

##### Severity Distribution Analysis

Severity distribution analysis will examine the frequency and proportional distribution of predicted NYHA and GOLD stages across datasets. Predicted severity distributions will be compared against clinically expected disease severity patterns within intensive care and hospital-based populations.

The analysis will help identify:

- stage imbalance,
- overclassification,
- underclassification,
- and potential dataset-specific bias.

Visualization techniques including heatmaps and severity distribution plots will be used to illustrate classification patterns across datasets and disease categories.

### Objective 3: Diagnostic Performance

The third objective will evaluate framework performance against confirmed HF and COPD diagnoses contained within clinical records.

#### Comparative Analysis

##### Sensitivity

Sensitivity will measure the framework’s ability to correctly identify true disease-related severity classifications.

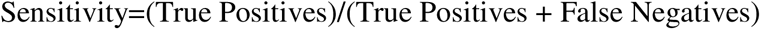

##### Specificity

Specificity will measure the framework’s ability to correctly exclude inappropriate disease classifications.

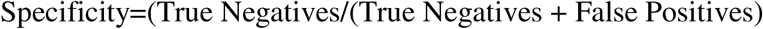

##### Positive Predictive Value (PPV)

PPV will evaluate the proportion of predicted positive classifications that are truly correct.

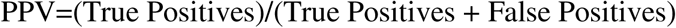

##### Negative Predictive Value (NPV)

NPV will assess the proportion of predicted negative classifications that are truly negative.

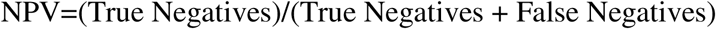

##### Confusion Matrices

Confusion matrices will be generated to visualize classification performance across severity categories. The matrices will illustrate:

- true positives,
- false positives,
- false negatives,
- and true negatives,

for both NYHA and GOLD severity predictions.

#### Correlation Analysis

##### Spearman Correlation

Spearman rank correlation analysis will assess monotonic relationships between predicted severity stages and severity confidence scores generated by the framework. Spearman correlation is appropriate because severity stages represent ordinal categories.

##### Pearson Correlation

Pearson correlation analysis will evaluate linear associations between continuous severity confidence scores and ordinal severity rankings derived from guideline-based classifications. Together, these analyses as summarized in Table 2 will provide comprehensive assessment of clinical validity, predictive consistency, and reliability of the ontology-integrated NLP framework across diverse clinical text repositories.

**Table 2.**
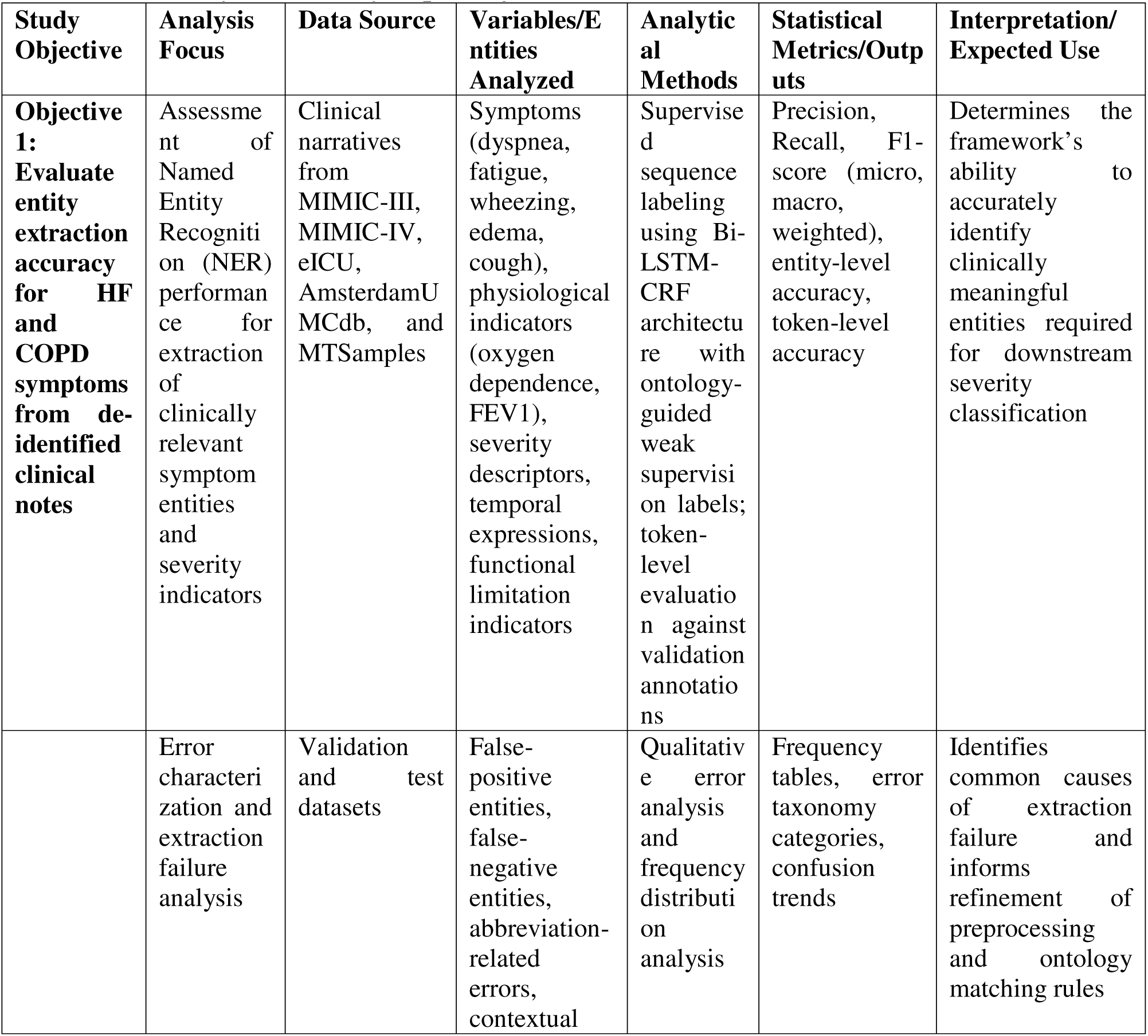
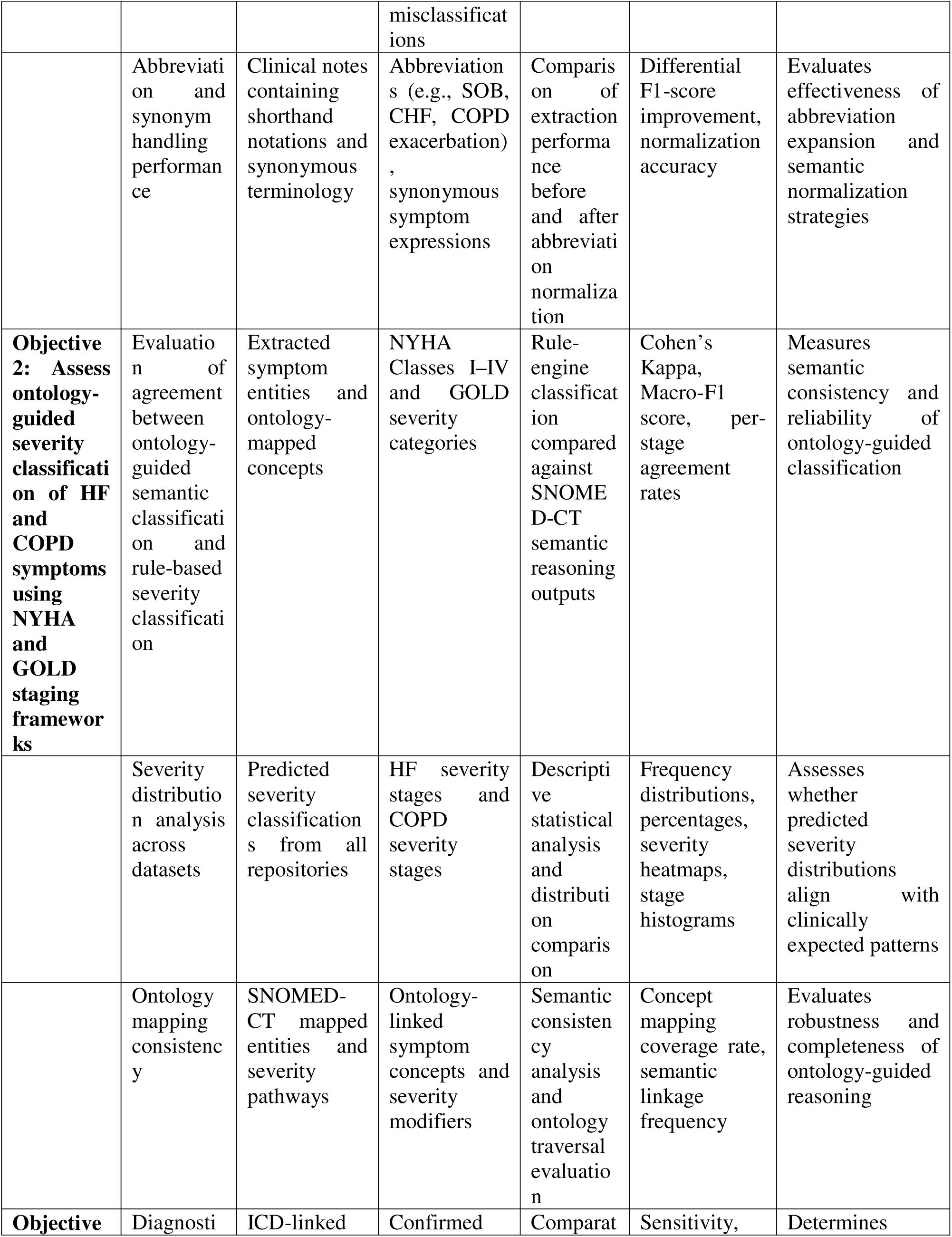
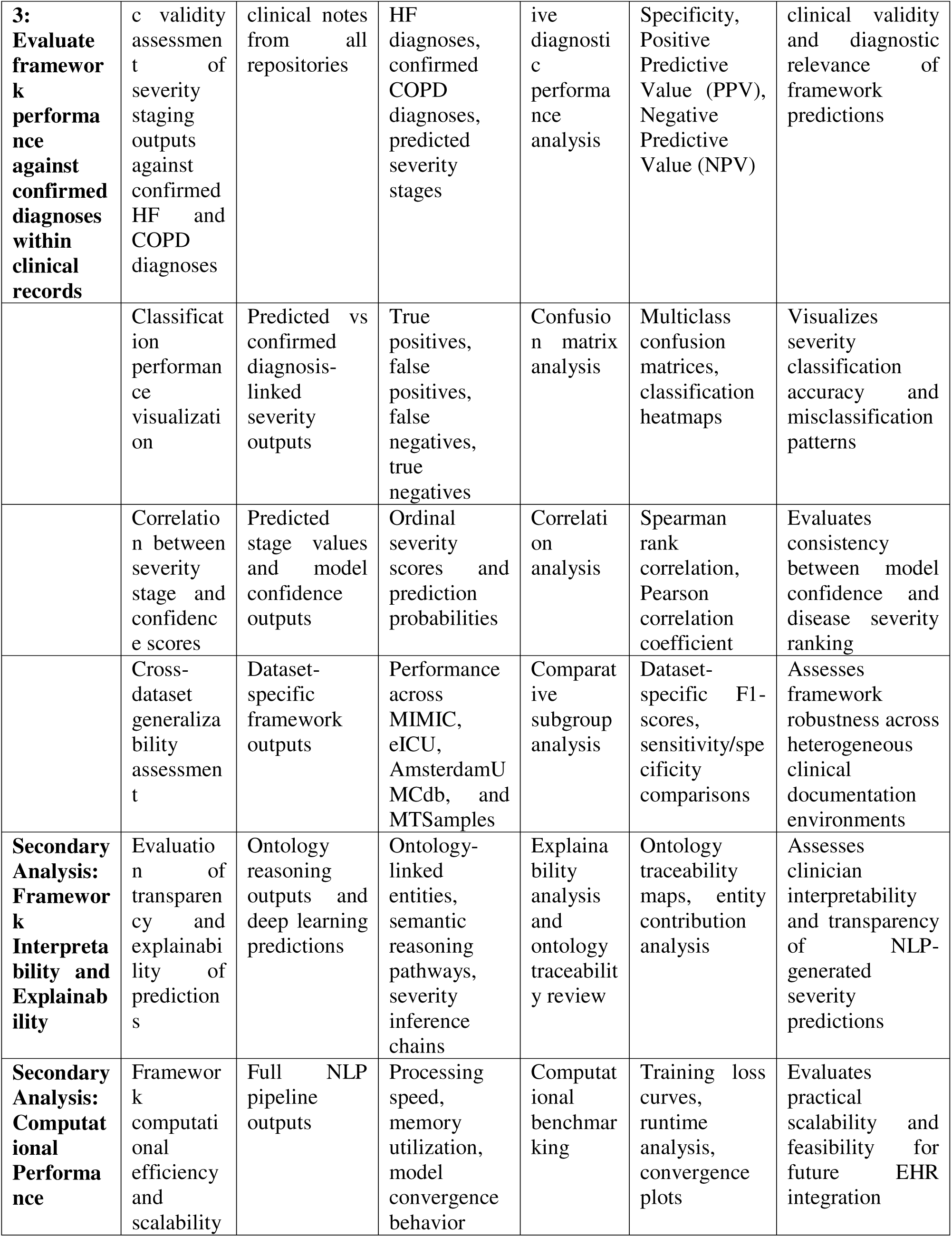
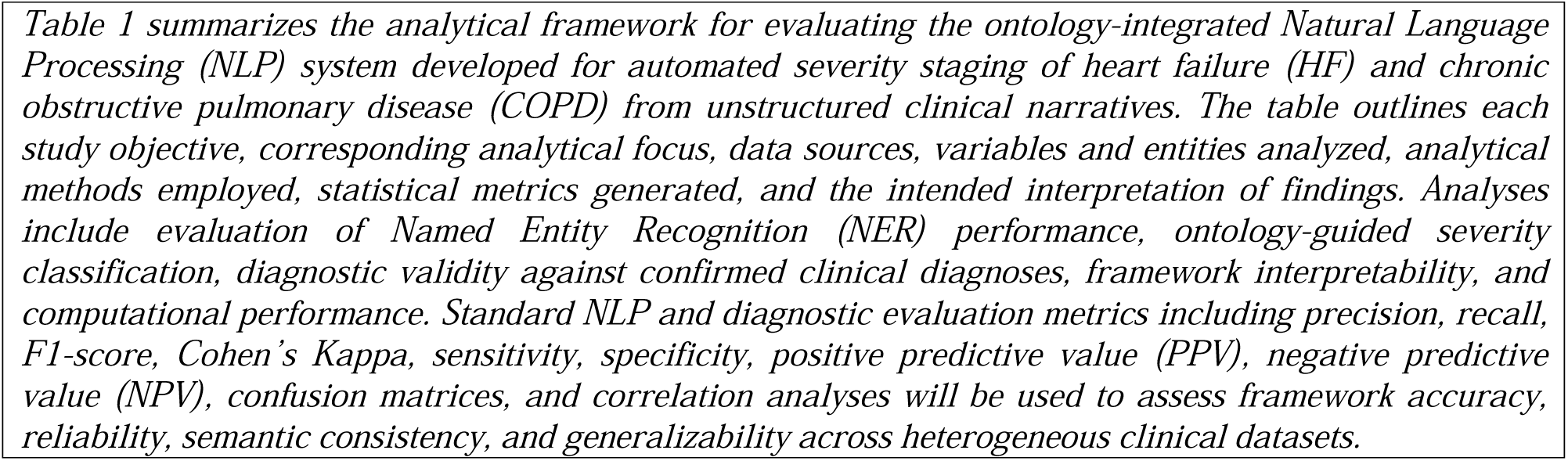
Summary of the analysis per objective.

#### Validation Strategy

##### Internal Validation

Internal validation will be conducted to evaluate the robustness, stability, and predictive performance of the ontology-integrated Natural Language Processing (NLP) framework within the sampled datasets. The complete dataset of de-identified clinical notes will be divided into training, validation, and testing subsets using a stratified 70:15:15 split to ensure balanced representation of heart failure (HF) and chronic obstructive pulmonary disease (COPD) records across all repositories.

The training dataset will be used for supervised learning and optimization of the Bidirectional Long Short-Term Memory with Conditional Random Field (Bi-LSTM-CRF) architecture, while the validation dataset will support hyperparameter tuning, early stopping, and optimization of model convergence. The testing dataset will be reserved exclusively for final model evaluation to minimize overfitting and ensure unbiased assessment of framework performance.

Internal validation will focus on:

- entity extraction accuracy,
- ontology-guided severity classification consistency,
- and diagnostic prediction performance.

Standard evaluation metrics including precision, recall, F1-score, sensitivity, specificity, positive predictive value (PPV), negative predictive value (NPV), and confusion matrices will be used to assess model stability and predictive reliability. Cross-validation approaches will additionally be explored during model tuning to improve generalizability and reduce variance associated with dataset partitioning.

##### Cross-dataset Validation

Cross-dataset validation will be performed to assess the generalizability and portability of the NLP framework across heterogeneous Electronic Health Record (EHR) environments. Because the study incorporates multiple publicly available repositories including MIMIC-III, MIMIC-IV, eICU Collaborative Research Database, AmsterdamUMCdb, and MTSamples, the framework will be evaluated across datasets with varying documentation styles, institutional practices, and clinical terminology.

Dataset-specific performance metrics will be generated to determine whether the framework maintains consistent entity extraction and severity classification performance across different clinical settings. Comparative analysis will assess:

- linguistic variability,
- institution-specific abbreviation patterns,
- note structure differences,
- and ontology mapping consistency.

Cross-dataset validation is important because many healthcare NLP systems demonstrate reduced performance when applied outside their original training environments due to differences in clinical language and documentation practices (Kersloot et al., 2020). Demonstrating stable performance across multiple repositories will strengthen evidence for the framework’s scalability and applicability in real-world healthcare systems.

##### Clinical Validity Assessment

Clinical validity assessment will evaluate the extent to which framework-generated severity classifications align with established clinical staging guidelines and confirmed diagnoses within the datasets. The framework outputs will be compared against:

- diagnosis-linked clinical documentation,
- ICD-9 and ICD-10 coded diagnoses,
- and expected severity distributions for HF and COPD populations.

For HF, predicted New York Heart Association (NYHA) stages will be evaluated against documented symptom burden and functional limitation descriptions. For COPD, predicted Global Initiative for Chronic Obstructive Lung Disease (GOLD) stages will be assessed using symptom narratives, oxygen dependence indicators, and available airflow limitation descriptors. Clinical validity assessment will also evaluate semantic coherence between extracted entities and predicted severity stages. Cases involving clinically implausible severity assignments will undergo targeted qualitative review to identify ontology reasoning failures, contextual ambiguity, or model misclassification patterns.

The integration of ontology-guided semantic reasoning is expected to improve explainability and clinical interpretability of framework outputs compared with black-box deep learning systems. Clinical validity therefore extends beyond predictive accuracy to include semantic consistency and transparency of severity inference pathways.

### Data Management

#### Data Storage

All study datasets will be stored within secure password-protected computing environments accessible only to authorized members of the research team. De-identified clinical datasets retrieved from publicly available repositories will be maintained in encrypted local storage systems and institutionally approved research workstations.

Structured outputs generated during preprocessing, ontology mapping, and deep learning stages will be stored separately from raw text datasets to improve data organization and facilitate reproducibility. Backup copies of datasets and analytical outputs will be maintained within encrypted external storage systems to minimize risk of accidental data loss.

#### Data Cleaning

Data cleaning procedures will be conducted prior to preprocessing and analysis to ensure consistency, quality, and integrity of the clinical narratives. Cleaning procedures will include:

- removal of duplicate records,
- normalization of malformed text,
- elimination of corrupted entries,
- handling of missing values,
- and standardization of encoding formats.

Clinical abbreviations and shorthand expressions will be normalized using domain-specific abbreviation dictionaries to improve downstream ontology matching and semantic consistency. Data quality checks will be conducted iteratively during preprocessing and model development to identify inconsistencies or processing failures.

#### Data Security

Data security measures will be implemented throughout the study to protect de-identified clinical datasets and analytical outputs. Access to data repositories requiring authentication, such as PhysioNet-hosted databases, will be restricted to certified study investigators who have completed mandatory data-use training requirements.

All datasets will remain de-identified, and no attempts will be made to re-identify individuals contained within the repositories. Secure access credentials, encrypted storage systems, and restricted user permissions will be employed to minimize unauthorized access risks.

Computational analyses involving cloud-based query environments such as Google BigQuery will be conducted through authenticated institutional accounts compliant with repository usage agreements and data governance standards.

#### Version Control

Version control procedures will be implemented to ensure reproducibility and traceability of framework development. All source code, preprocessing scripts, ontology mapping procedures, and analytical workflows will be maintained using Git-based version control systems.

Version tracking will document:

- code modifications,
- model architecture updates,
- hyperparameter adjustments,
- ontology dictionary refinements,
- and preprocessing pipeline changes.

This approach will support collaborative development, reproducibility of findings, and transparent documentation of methodological evolution throughout the study.

#### GitHub/Open-source Release Plan

Following publication and completion of the study, the NLP framework source code, preprocessing scripts, ontology integration workflows, and analytical pipelines will be released through a publicly accessible GitHub repository. The open-source release will support:

- reproducibility,
- transparency,
- external validation,
- and future adaptation of the framework in healthcare NLP research.

The released repository will include:

- implementation documentation,
- dependency specifications,
- installation guidelines,
- preprocessing workflows,
- and model evaluation scripts.

No raw clinical text or protected repository data will be publicly shared in compliance with repository usage agreements and ethical data governance standards.

## Ethical Considerations

### Data Access Permissions

#### PhysioNet

Access to MIMIC-III, MIMIC-IV, and the eICU Collaborative Research Database will be obtained through PhysioNet after successful completion of mandatory Collaborative Institutional Training Initiative (CITI) data security and human research ethics training. Data use agreements governing access, storage, and analysis of de-identified datasets will be strictly observed throughout the study.

#### AmsterdamUMCdb

Access to AmsterdamUMCdb will be obtained through compliance with the repository’s publicly available access procedures and data-use policies. All analytical activities involving AmsterdamUMCdb data will adhere to institutional and repository-specific governance requirements.

#### MTSamples Terms of Use

The MTSamples repository will be used in accordance with publicly available terms of use governing educational and research utilization of medical transcription samples. Data extraction procedures will be limited to publicly accessible clinical narratives and will not involve collection of identifiable patient information.

#### Confidentiality and Security

Although the study utilizes publicly available de-identified datasets, confidentiality and security principles will be maintained throughout all phases of data handling and analysis. No patient-identifiable information will be collected, stored, analyzed, or published.

All outputs generated from the framework will contain only anonymized identifiers and derived severity classifications. Research findings will be presented in aggregate form without disclosure of identifiable institutional or patient-specific information.

#### Use of De-identified Data

The study exclusively utilizes secondary de-identified clinical datasets that have undergone formal anonymization procedures before public release. Because no direct patient interaction or identifiable information is involved, the study presents minimal risk to individuals represented within the datasets.

No attempts will be made to re-identify individuals or link clinical records to external identifiable data sources. Use of de-identified datasets substantially reduces ethical risks associated with privacy breaches and confidentiality violations.

#### Ethical Compliance

The study will comply with institutional research ethics standards, repository-specific data governance policies, and international principles governing ethical artificial intelligence research. Repository access conditions, licensing agreements, and data-use restrictions will be strictly observed.

The framework development process will additionally adhere to principles of responsible artificial intelligence including:

- transparency,
- fairness,
- reproducibility,
- accountability,
- and explainability.

All analytical procedures will be conducted solely for scientific and educational purposes.

#### AI Transparency and Explainability

Artificial intelligence transparency and explainability are central ethical considerations within healthcare NLP research. The proposed framework integrates ontology-guided semantic reasoning with deep learning approaches to improve interpretability of severity predictions and reduce reliance on opaque black-box models.

Explainability mechanisms within the framework will allow tracing of:

- extracted symptom entities,
- ontology mappings,
- severity weighting pathways,
- and rule-based inference logic.

These mechanisms are intended to improve clinician trust, facilitate reproducibility, and support future integration into clinical decision-support systems. Transparent reasoning pathways are particularly important in healthcare environments where AI-generated outputs may influence patient care, risk stratification, and clinical decision-making.

#### Expected Outcomes and Significance

##### Scientific Contributions

This study is expected to contribute significantly to the growing field of clinical Natural Language Processing (NLP) by developing and evaluating a hybrid ontology-integrated framework for automated severity staging of heart failure (HF) and chronic obstructive pulmonary disease (COPD). The framework will advance current healthcare NLP research by integrating symbolic ontology-based reasoning with deep learning sequence labeling techniques to support clinically meaningful severity classification from unstructured Electronic Health Record (EHR) narratives.

The study is anticipated to contribute methodological innovations in:

- ontology-guided weak supervision,
- hybrid rule-based and deep learning architectures,
- semantic normalization of clinical narratives,
- and explainable severity inference mechanisms.

In addition, the study will provide empirical evidence regarding the feasibility of automated NYHA and GOLD severity staging without reliance on manually annotated severity corpora. This contribution is particularly important because annotated clinical datasets remain scarce in many healthcare systems.

The proposed framework may further provide a reusable methodological foundation for future ontology-integrated NLP applications involving:

- disease phenotyping,
- clinical risk prediction,
- retrospective clinical audit,
- and automated EHR analytics.

##### Clinical Relevance

The framework has potential clinical relevance because severity staging is central to diagnosis, treatment planning, hospitalization decisions, and longitudinal monitoring of HF and COPD patients. Automated extraction and classification of symptom severity from clinical narratives may support clinicians by improving identification of disease progression and enhancing access to structured severity information within EHR systems.

The framework may additionally improve:

- retrospective patient stratification,
- clinical audit processes,
- healthcare quality monitoring,
- and disease surveillance activities.

By converting free-text clinical narratives into structured severity classifications, the framework may reduce dependence on incomplete structured coding systems and improve availability of clinically actionable information for healthcare providers and researchers.

The integration of explainable ontology-guided reasoning may also improve clinician trust and facilitate future incorporation into clinical decision-support systems.

##### Health Informatics Implications

This study is expected to contribute to broader health informatics initiatives involving semantic interoperability, artificial intelligence integration, and secondary use of EHR data. Clinical narratives contain substantial unstructured information that remains underutilized in healthcare systems due to challenges in automated interpretation. The proposed framework addresses this gap by enabling structured extraction of symptom severity information from free-text records.

Ontology integration using SNOMED-CT is anticipated to improve interoperability between healthcare systems by standardizing heterogeneous clinical terminology into semantically consistent representations. This may support:

- harmonized clinical analytics,
- population health surveillance,
- data integration across institutions,
- and interoperability of AI-enabled healthcare systems.

The framework may further contribute to development of explainable and transparent healthcare AI systems by combining neural sequence learning with interpretable ontology-guided semantic reasoning pathways.

##### Applicability in Resource-Constrained Settings

The proposed framework may have important applications in resource-constrained healthcare environments where structured EHR systems and manually annotated datasets are limited. Many low- and middle-income countries possess increasing volumes of unstructured clinical documentation but limited infrastructure for advanced healthcare analytics.

Because the framework relies partly on ontology-guided weak supervision rather than extensive manually annotated corpora, it may provide a scalable and cost-effective approach for extracting clinically relevant severity information from routine clinical documentation. The use of publicly available datasets and open-source implementation strategies may further improve adaptability in low-resource research and healthcare settings.

In addition, automated severity staging may support:

- disease surveillance,
- retrospective audit,
- clinical prioritization,
- and healthcare planning,

particularly in environments with constrained specialist capacity and limited access to advanced clinical decision-support infrastructure.

## Discussion

### Anticipated Contributions

This study is expected to demonstrate the feasibility of integrating ontology-guided semantic reasoning with deep learning approaches for automated severity staging of HF and COPD from unstructured clinical narratives [30]. The hybrid framework is anticipated to address several limitations associated with conventional healthcare NLP systems, particularly dependence on manually annotated datasets and inadequate clinical interpretability [40].

The anticipated findings may contribute to the growing body of evidence supporting hybrid AI architectures that combine symbolic reasoning and statistical learning within healthcare applications. The study may further demonstrate the utility of ontology-guided weak supervision strategies in clinical NLP environments characterized by limited annotated data availability[41]. The framework is also expected to contribute methodological insights into; semantic normalization, ontology-assisted sequence labeling, and explainable clinical severity classification [42].

### Potential Clinical Utility

The proposed framework may have practical utility within clinical and health systems environments by enabling automated extraction of clinically meaningful severity information from routine EHR documentation. Such functionality may support: clinical audit, patient stratification, disease monitoring, and retrospective outcomes research [43].

The framework may further assist healthcare institutions in identifying high-risk patients through automated severity classification without requiring extensive manual chart review. This may improve efficiency of population-level disease surveillance and facilitate more timely access to clinically relevant information [44].

Although the framework is not intended for immediate real-time deployment, the findings may provide a foundation for future development of clinical decision-support systems capable of integrating severity inference into routine healthcare workflows.

### Role of Ontology Integration

Ontology integration represents a central component of the proposed framework because clinical narratives frequently contain heterogeneous terminology, abbreviations, synonymous expressions, and institution-specific language patterns. By integrating SNOMED-CT semantic mappings, the framework is expected to improve consistency of clinical entity normalization and reduce ambiguity associated with free-text documentation [45].

Ontology-guided reasoning may additionally enhance framework coverage by enabling severity inference even when explicit NYHA or GOLD classifications are absent from clinical notes. The semantic relationships encoded within SNOMED-CT may allow the framework to identify clinically meaningful associations between symptoms, physiological findings, and severity indicators [15].

Furthermore, ontology integration is expected to improve semantic interoperability and facilitate reproducibility of NLP outputs across heterogeneous clinical datasets.

### Explainability and Interoperability

Explainability remains a major concern in healthcare artificial intelligence because opaque “black-box” systems may limit clinician trust and reduce adoption within clinical settings. The proposed framework addresses this challenge by incorporating interpretable ontology-guided reasoning alongside deep learning sequence models [46].

The framework is expected to provide traceable severity inference pathways linking: extracted symptom entities, ontology mappings, severity weighting mechanisms, and final classification outputs.

This transparency may improve interpretability and support future integration into clinical decision-support systems.

Interoperability is also expected to improve through semantic normalization using standardized SNOMED-CT terminology. This may facilitate integration of framework outputs across healthcare systems and support broader health informatics applications involving structured secondary use of EHR narratives [47].

### Limitations

This study has several anticipated limitations. First, the framework depends entirely on secondary publicly available datasets, which may contain inconsistencies, incomplete documentation, or institution-specific language patterns that could influence model performance. Second, substantial heterogeneity exists across the selected datasets with respect to note structure, terminology, formatting, and documentation practices. Although this diversity supports evaluation of generalizability, it may also introduce variability that affects classification consistency.

Third, external validation beyond the selected repositories will be limited. The framework will not be evaluated using prospectively collected institutional datasets or real-world deployment environments, which may limit immediate generalizability to clinical practice settings.

Fourth, the framework will process only English-language clinical narratives. Consequently, the findings may not generalize to multilingual healthcare environments or non-English clinical documentation systems.

Finally, the study does not involve real-time clinical deployment or prospective integration into live EHR systems. Therefore, workflow integration challenges, clinician interaction dynamics, and operational implementation barriers will not be assessed within the current protocol.

### Dissemination Plan

Findings from this study will be disseminated through multiple scientific and open-science platforms to maximize accessibility, transparency, and reproducibility.

The study protocol and final findings will be submitted for publication in peer-reviewed journals. Research findings will additionally be presented at local and international scientific conferences related to: biomedical informatics, artificial intelligence, digital health, and clinical decision support systems.

To promote transparency and reproducibility, the framework source code, preprocessing scripts, ontology integration workflows, and analytical pipelines will be released through a publicly accessible GitHub repository following publication. The repository will include: technical documentation, installation instructions, dependency specifications, and workflow implementation guidelines.

No raw clinical text or restricted repository data will be publicly shared in compliance with repository governance and data-use agreements.

## Data Availability

All data produced in the present study are available upon reasonable request to the authors

## Authors’ Contributions

JI conceptualized the study, developed the methodological framework, designed the NLP architecture, and drafted the manuscript. FMM contributed to methodological refinement, computational framework development, and critical review of the manuscript. RO contributed to ontology integration strategy, analytical framework development, and critical revision of the manuscript. All authors reviewed and approved the final manuscript. MSS contributed to study design, clinical interpretation, health informatics perspectives, and manuscript review.

## Funding Statement

This study has not received external funding at the time of protocol development. The research will be conducted using publicly available datasets and institutional computational resources. Any future funding support obtained during the course of the study will be appropriately disclosed in subsequent publications.

## Competing Interests

The authors declare that they have no competing interests related to this study.

## Data Availability Statement

The datasets used in this study are publicly available through their respective repositories subject to compliance with repository-specific access requirements and data-use agreements. MIMIC-III, MIMIC-IV, and eICU Collaborative Research Database are accessible through PhysioNet following completion of required training and credentialing procedures. AmsterdamUMCdb is accessible through the official repository under applicable usage policies. MTSamples clinical narratives are publicly accessible for educational and research purposes.

The source code, preprocessing scripts, ontology integration workflows, and analytical pipelines developed during this study will be made publicly available through a GitHub repository upon publication of the study findings.

## Notes

### Competing Interest Statement

The authors have declared no competing interest.

### Author Declarations

Approximately 1,000 de-identified clinical notes will be sampled proportionately from publicly available repositories including MIMIC-III/IV, eICU Collaborative Research Database, AmsterdamUMCdb, and MTSamples. Clinical text preprocessing will include tokenization, lemmatization, dependency parsing, abbreviation expansion, and negation detection. Ontology-guided semantic normalization will map extracted symptom entities to standardized SNOMED-CT concepts to support severity staging.

